# Prevalence, intensity, and risk factors of Schistosomiasis and Intestinal Parasitic Infections among primary school children in northern Uganda: Implications for public health interventions

**DOI:** 10.1101/2025.02.13.25322200

**Authors:** John Paul Byagamy, Robert Opiro, Margaret Nyafwono, Geoffrey Maxwell Malinga, Richard Echodu, Emmanuel Igwaro Odongo-Aginya

## Abstract

**Background:** Schistosomiasis and intestinal parasitic infections are major public health challenges in Uganda, particularly among school-aged children. These infections contribute to malnutrition, anemia, and impaired cognitive development, limiting children’s educational and overall well-being. The Lango sub-region in Northern Uganda is endemic for *Schistosoma mansoni*, but limited epidemiological data exist to inform targeted control strategies. This study aimed to determine the prevalence, intensity, and risk factors associated with schistosomiasis and intestinal parasitic infections among primary school children in the region.

**Methodology/Principal Findings:** We conducted a cross-sectional study between January and March 2023, involving 802 primary school children from randomly selected schools in Lira District, Lira City, and Kole District. Stool and urine samples were examined using the Odongo-Aginya method, urine filtration, and Point-of-Care Circulating Cathodic Antigen (POC-CCA) test to detect *S. mansoni*, *S. haematobium*, and intestinal parasites. Data on potential risk factors were collected via structured interviews and analyzed using logistic regression in SPSS version 25.0.

The overall prevalence of schistosomiasis was 34.5% (*S. mansoni*), with light (11.6%), moderate (5.4%), and heavy (2.9%) infection intensities. Other intestinal parasites were detected in 20.3% of participants, including *Ascaris lumbricoides* (11.6%) and hookworms (6.4%). Key risk factors for schistosomiasis included bathing with borehole water (OR = 6.017, p = 0.019) and low paternal education level (p = 0.05), while recent praziquantel treatment was protective (OR = 0.122, p = 0.009).

**Conclusions/Significance:** Our findings highlight a high burden of schistosomiasis and intestinal parasitic infections in Northern Uganda, emphasizing the need for strengthened mass drug administration (MDA), improved water, sanitation, and hygiene (WASH) infrastructure, and targeted health education. Addressing these factors through integrated, community-driven interventions is essential for sustainable disease control and prevention.

**Author Summary:** Schistosomiasis and intestinal parasitic infections are significant public health challenges, especially in low-resource settings like Northern Uganda. These infections can lead to malnutrition, anemia, and impaired cognitive development, particularly in school-aged children. In this study, we investigated the prevalence, intensity, and risk factors for these infections among primary school children in the Lango sub-region of Northern Uganda.

We examined stool and urine samples from over 800 children and found that more than one-third were infected with *Schistosoma mansoni*, the parasite responsible for intestinal schistosomiasis. Additionally, we identified other intestinal parasites, such as *Ascaris lumbricoides* (roundworms) and *hookworms*. Our findings show that factors like bathing with borehole water, low parental education levels, and district of residence were linked to a higher risk of infection. However, children who had recently received treatment with praziquantel, the main drug used for schistosomiasis, were less likely to be infected.

These results highlight the urgent need for expanded treatment programs, improved sanitation, and targeted health education efforts to reduce the burden of parasitic infections in Ugandan schoolchildren. By identifying key risk factors, our study provides valuable insights to guide policymakers and public health professionals in designing more effective intervention strategies.

## Introduction

Schistosomiasis and intestinal parasitic infections are among the most prevalent neglected tropical diseases globally [1], disproportionately affecting populations in low- and middle-income countries [2]. These infections, caused by trematodes and soil-transmitted helminths, respectively, contribute significantly to morbidity, particularly in children, due to their impact on growth, cognitive development, and overall health [3, 4]. In sub-Saharan Africa, where sanitation and access to clean water are limited, schistosomiasis and intestinal parasitic infections remain pervasive public health challenges [5–7].

In Uganda, *Schistosoma mansoni* is endemic in many regions, including the Lango sub-region in Northern Uganda [8, 9]. However, it’s only in the Lango sub-region known to harbor both *Schistosoma mansoni* and *Schistosoma haematobium* [10–12]. The disease is primarily transmitted through contact with freshwater bodies contaminated with cercariae released by infected snail hosts [13, 14]. Children are especially vulnerable due to frequent water contact activities, such as swimming, fishing, and fetching water, often in infested environments [15–17]. Beyond schistosomiasis, soil-transmitted helminths, including *Ascaris lumbricoides*, *Hookworms*, and *Trichuris trichiura*, are widespread, perpetuating cycles of poverty and illness [18, 19].

Efforts to control these infections in Uganda include mass drug administration (MDA) campaigns using Praziquantel and Albendazole, supported by the Ministry of Health and international partners [20, 21]. However, persistent transmission, coupled with uneven intervention coverage and compliance, has limited progress. Environmental factors, socio-economic disparities, and lack of community-level data further hinder effective control strategies [22, 23].

Several studies have reported varying prevalence rates of schistosomiasis and intestinal parasitic infections across Uganda [8, 24, 25]. For instance, the prevalence of *Schistosoma mansoni* in school-aged children ranges from 20% to 60%, depending on the region [15, 26]. However, there is a paucity of data specific to the Lango sub-region, where unique environmental and socio-economic conditions may influence disease distribution and risk factors [18, 27].

This study aimed to bridge the knowledge gap by determining the prevalence, intensity, and risk factors associated with schistosomiasis and intestinal parasitic infections among primary school children in the Lango sub-region. The findings aim to inform local and national policy-makers and stakeholders on targeted intervention strategies to reduce the disease burden in Northern Uganda.

## Methods and Materials

### Study Area

This study was conducted in the Lango sub-region, Northern Uganda, specifically targeting three administrative areas: Lira District, Lira City, and Kole District[28]. These districts are among the nine districts in the Lango sub-region, predominately inhibited by the Lango tribe. The main economic activities are commercial and subsistence farming, small-scale fishing, retail, and wholesale business. These regions are characterized by a tropical climate, seasonal rainfall, and reliance on freshwater sources for domestic and recreational activities, which predispose residents to schistosomiasis transmission[29].

**Figure 1:**
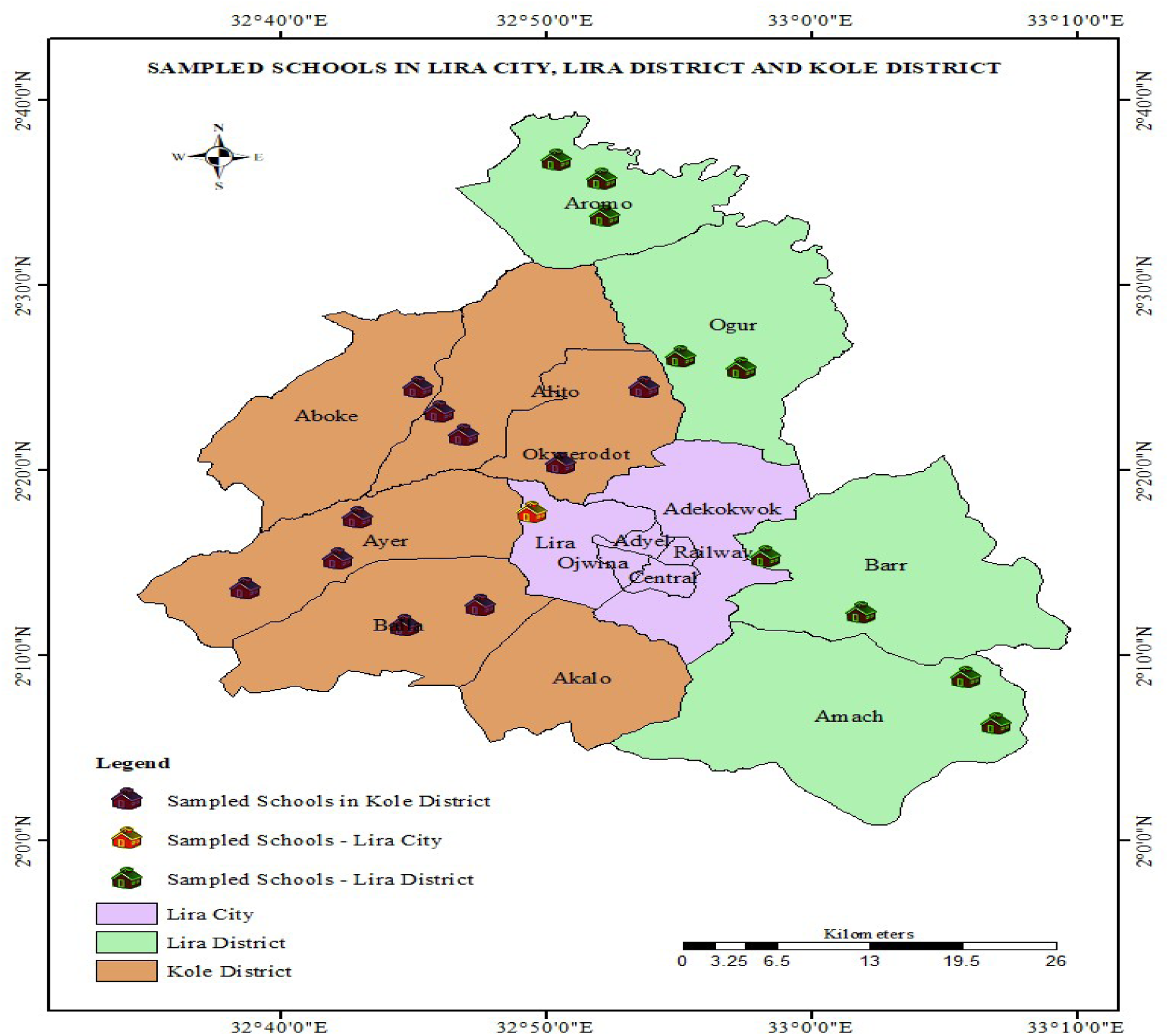
Study sites showing schools sampled

### Study Design

A cross-sectional study design was employed to assess the prevalence, intensity, and risk factors associated with schistosomiasis and intestinal parasitic infections among school-aged children. Data collection occurred between January and March 2023.

### Study Population

The study targeted primary school children aged 5–16 years enrolled in Primary 1 to 7. This population was chosen because school-aged children are most at risk of schistosomiasis and intestinal parasitic infections due to frequent water contact and inadequate hygiene practices.

### Inclusion Criteria

The children aged 5–16 years attending selected primary schools in Lira District, Lira City, and Kole District, and at least lived there for at least one year, written informed consent obtained from parents or guardians for minors, and assent from children aged 8 years and above.

### Sample Size Determination

The sample size was calculated using the formula for cross-sectional studies [30]. Where: N = Z^2^×PQ/d^2.^ Where, N =the desired sample size, P = estimated prevalence, estimated to be 50%, Q = percentage of people not infected (1-p), d = degree of precision required, usually set at 0.05, Z = confidence limit at 95% interval (1.96). Adjusting for a 10% non-response rate, the final sample size was 802 participants.

### Sampling Methods

A multistage sampling technique was used. First, 20 primary schools were randomly selected from P1-P7 in the three districts. Within each school, participants were stratified by grade, and simple random sampling was employed to select children proportionate to the school population.

### Stool and Urine Examinations

Stool samples were collected in sterile containers and examined using the Odongo-Aginya method to detect intestinal parasites and quantify eggs per gram (epg) for *Schistosoma mansoni*. Meanwhile, urine samples were screened for *Schistosoma mansoni* antigens using the Point-of-Care Circulating Cathodic Antigen (POC-CCA) test. Urine filtration was used to confirm the absence of *Schistosoma haematobium*.

### Risk Factors and Demographic Data Collection

Data on potential risk factors, including water contact activities, sanitation practices, parental education, and prior praziquantel treatment, were collected through structured interviews. Demographic data, such as age, gender, and school, were recorded.

### Data Analysis

Data were analyzed using SPSS version 25.0 [31]. Descriptive statistics were used to calculate the prevalence and intensity of infections. The intensity of *S. mansoni* infection was calculated using WHO guidelines [34], which classify infections into light, moderate, and heavy classes. Associations between risk factors and infection status were evaluated using chi-square tests, while logistic regression identified independent predictors of infection. Odds ratios (OR) with 95% confidence intervals (CI) were reported. Significance was set at 0.05.

## Results

### Sociodemographic Characteristics of Study Participants

The study included participants from three districts in the Lango sub-region. The highest proportion of participants came from Kole District 398 (49.6%), followed by Lira District 361 (45%), with a smaller representation from Lira City 43 (5.4%).

Gender distribution showed a higher percentage of males 441 (55%) than females 361 (45%). The mean age was 12.25, SD ± 2.113. The majority of participants were aged between 11-13 years 421 (52.5%), followed by those aged 14-16 years 229 (28.6%). Smaller proportions were observed in the 8-10 years 129 (16.1%) and 5-7 years 23 (2.9%) age groups (Table 1).

**Table 1:**
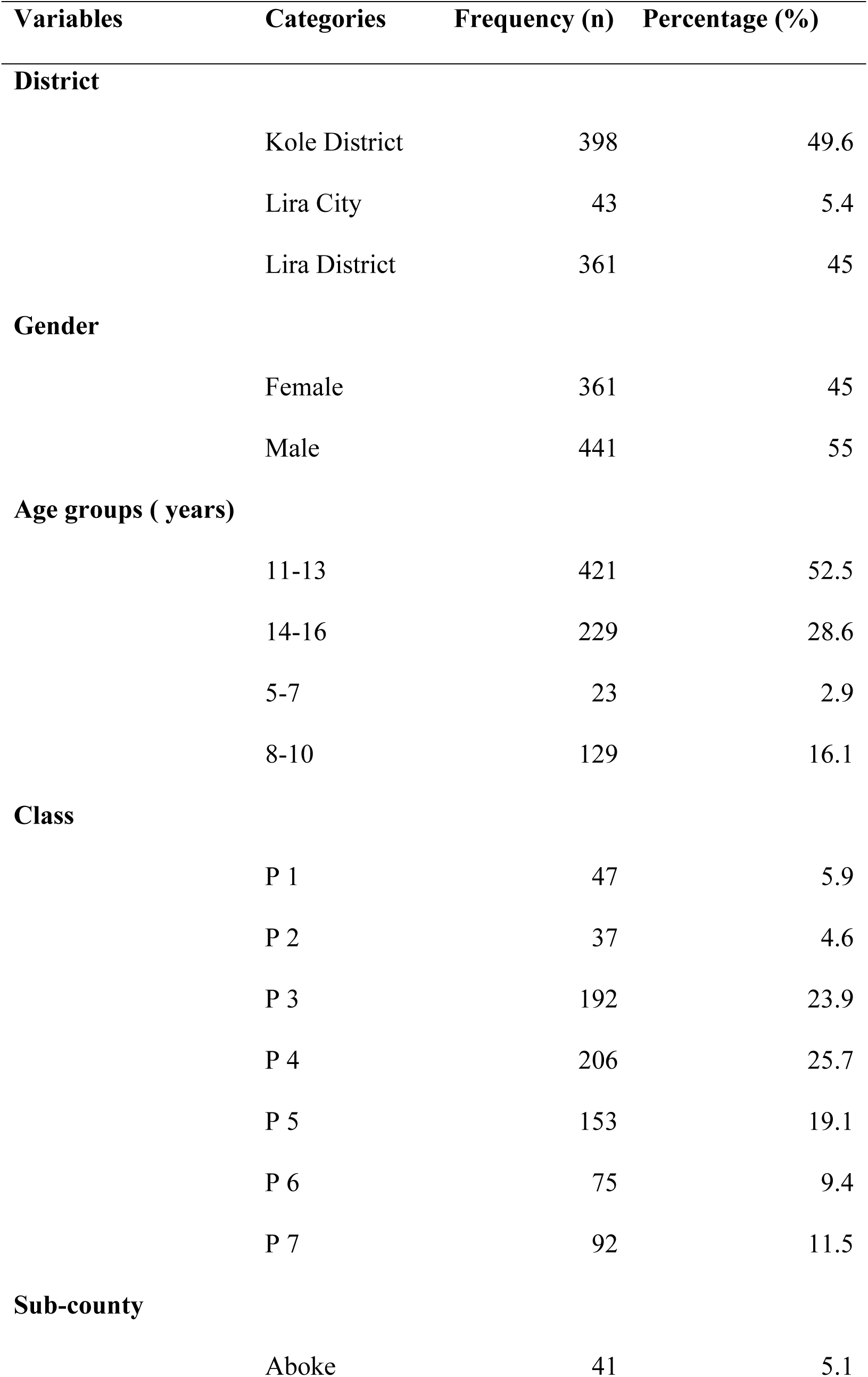
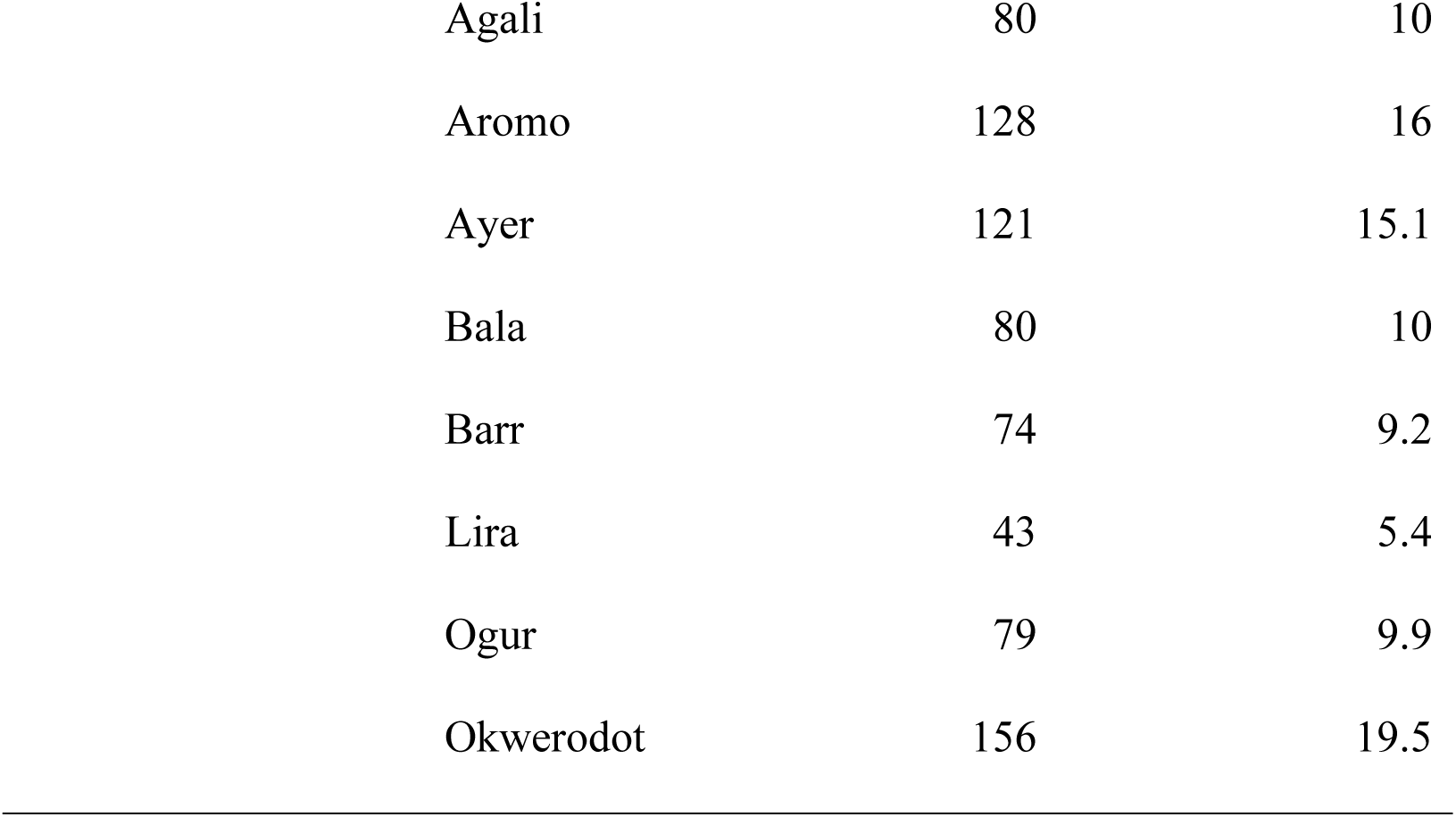
Sociodemographic characteristics of study participants in the Lango sub-region.

| **Variables** | **Categories** | **Frequency (n)** | **Percentage (%)** |
| --- | --- | --- | --- |
| **District** |  |  |  |
|  | Kole District | 398 | 49.6 |
|  | Lira City | 43 | 5.4 |
|  | Lira District | 361 | 45 |
| **Gender** |  |  |  |
|  | Female | 361 | 45 |
|  | Male | 441 | 55 |
| **Age groups ( years)** |  |  |  |
|  | 11-13 | 421 | 52.5 |
|  | 14-16 | 229 | 28.6 |
|  | 5-7 | 23 | 2.9 |
|  | 8-10 | 129 | 16.1 |
| **Class** |  |  |  |
|  | P 1 | 47 | 5.9 |
|  | P 2 | 37 | 4.6 |
|  | P 3 | 192 | 23.9 |
|  | P 4 | 206 | 25.7 |
|  | P 5 | 153 | 19.1 |
|  | P 6 | 75 | 9.4 |
|  | P 7 | 92 | 11.5 |
| **Sub-county** |  |  |  |
|  | Aboke | 41 | 5.1 |

|  |  |  |
| --- | --- | --- |
| Agali | 80 | 10 |
| Aromo | 128 | 16 |
| Ayer | 121 | 15.1 |
| Bala | 80 | 10 |
| Barr | 74 | 9.2 |
| Lira | 43 | 5.4 |
| Ogur | 79 | 9.9 |
| Okwerodot | 156 | 19.5 |

Participants were distributed across different primary school classes, with the largest groups in P4 206 (25.7%) and P3 192 (23.9%). Smaller percentages were seen in classes P1 47 (5.9%) and P2 37 (4.6%), while P5, P6, and P7 accounted for 153 (19.1%), 75 (9.4%), and 92 (11.5%), respectively (Table 1).

The sub-county distribution showed the highest representation from Okwerodot 156 (19.5%), followed by Aromo 128 (16%) and Ayer 121 (15.1%). Other sub-counties had lower proportions, such as Aboke 41 (5.1%) and Lira 43 (5.4%), Table 1.

### Overall Prevalence of Parasites Identified

The study identified various parasitic infections among participants, with *Schistosoma mansoni* being the most prevalent, affecting 277 (34.5%) of the participants. Other notable parasites included *Entamoeba coli* 94 (11.7%) and *Ascaris lumbricoides* 93 (11.6%).

Lower prevalence rates were observed for *Hookworms* 51 (6.4%), *Enterobius vermicularis* 26 (3.2%), and *Trichuris trichura* 12 (1.5%). Rare cases were recorded for *Fasciola species* 11 (1.4%), *Entamoeba histolytica* 10 (1.2%), and *Taenia species* 07 (0.9%).

Very low prevalence 01 (0.1%) was found for *Giardia lamblia*, *Paragonimus westermani*, *Strongyloides stercolaris*, and *Blastocystis hominis* (Table 2).

**Table 2:** overall prevalence of parasites identified.

| Parasites | No. Positive (n) | Prevalence (%) |
| --- | --- | --- |
| *Schistosoma mansoni* | 277 | 34.5 |
| *Ascaris lumbricoides* | 93 | 11.6 |
| *Hookworms* | 51 | 6.4 |
| *Trichuris trichura* | 12 | 1.5 |
| *Entamoeba coli* | 94 | 11.7 |
| *Entamoeba histolytica* | 10 | 1.2 |
| *Enterobius vermicularis* | 26 | 3.2 |
| *Fasciola species* | 11 | 1.4 |
| *Giardia lamblia* | 1 | 0.1 |
| *Paragonimus westermani* | 1 | 0.1 |
| *Strongyloides stercoralis* | 1 | 0.1 |
| *Taenia species* | 7 | 0.9 |
| *Blastocystis hominis* | 1 | 0.1 |

### Prevalence of *Schistosoma mansoni* among school children

The study evaluated the prevalence of *Schistosoma mansoni* infection based on sociodemographic, educational, and behavioral variables. The prevalence was highest in Lira District 159 (57.4%) and lowest in Lira City 08 (2.9%), with Kole District showing 110 (39.7%) positivity (p < 0.001). No significant difference was observed between males 158 (57.0%) and females 119 (43.0%) (p=0.396), (Table 3).

**Table 3:**
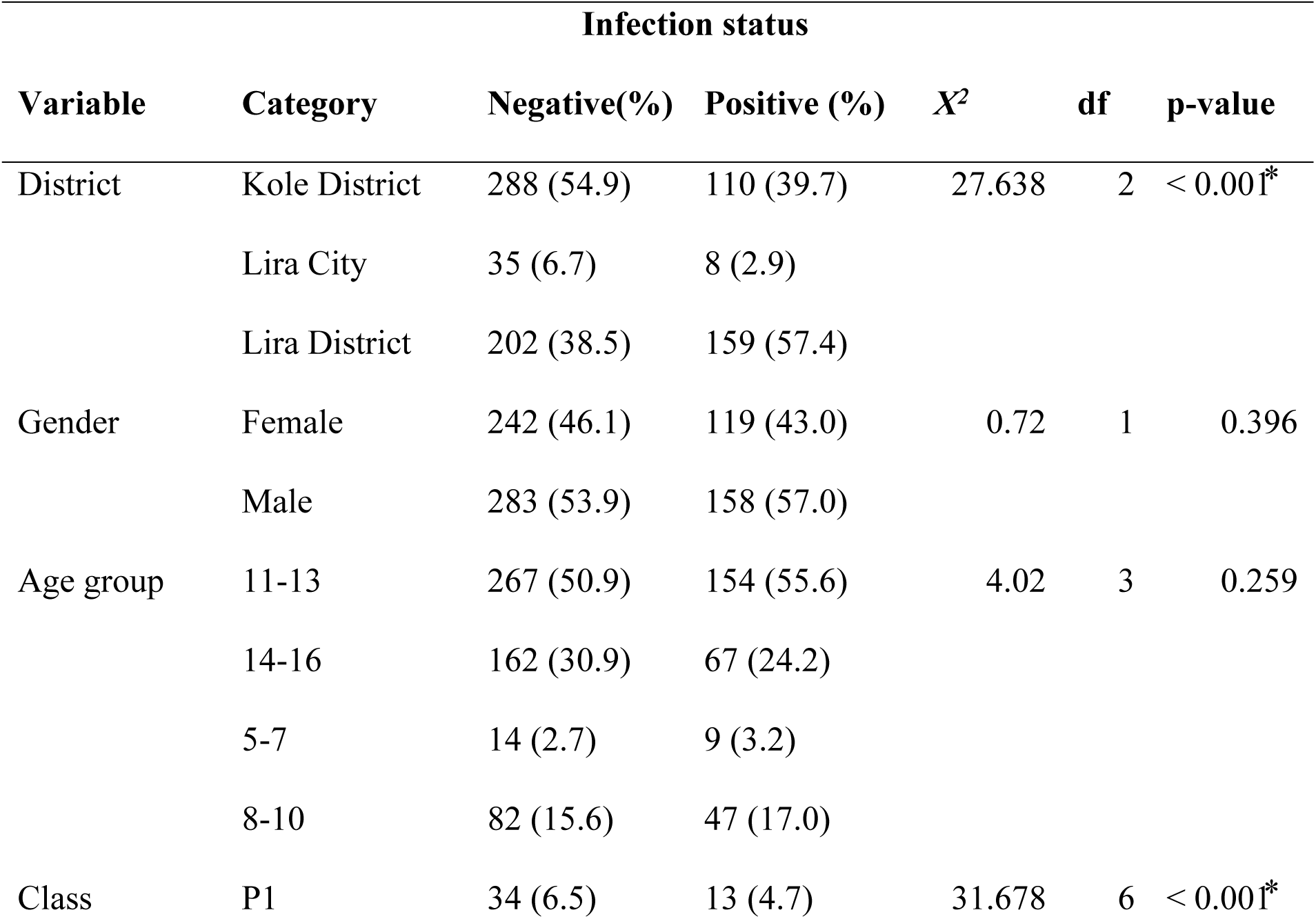
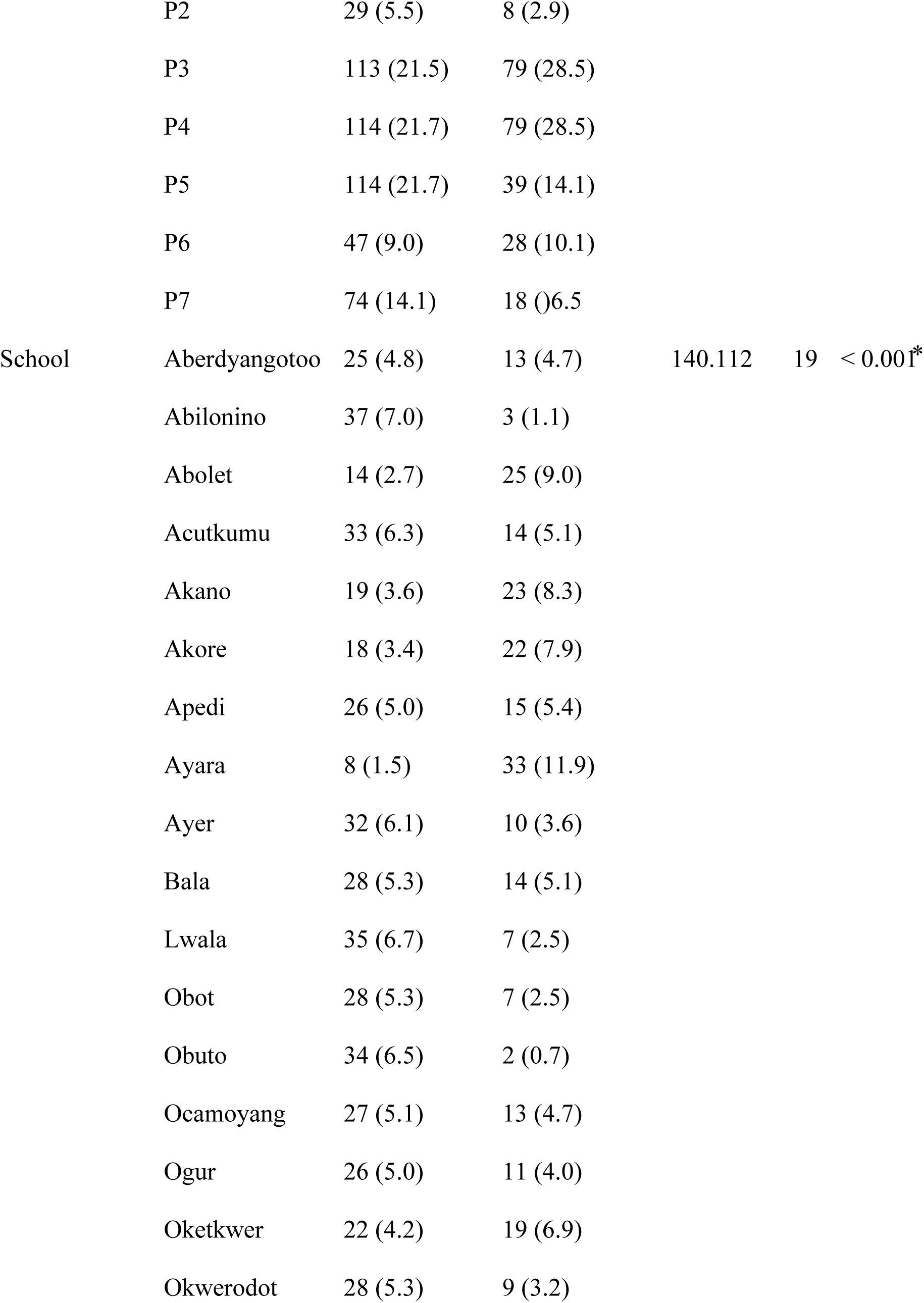
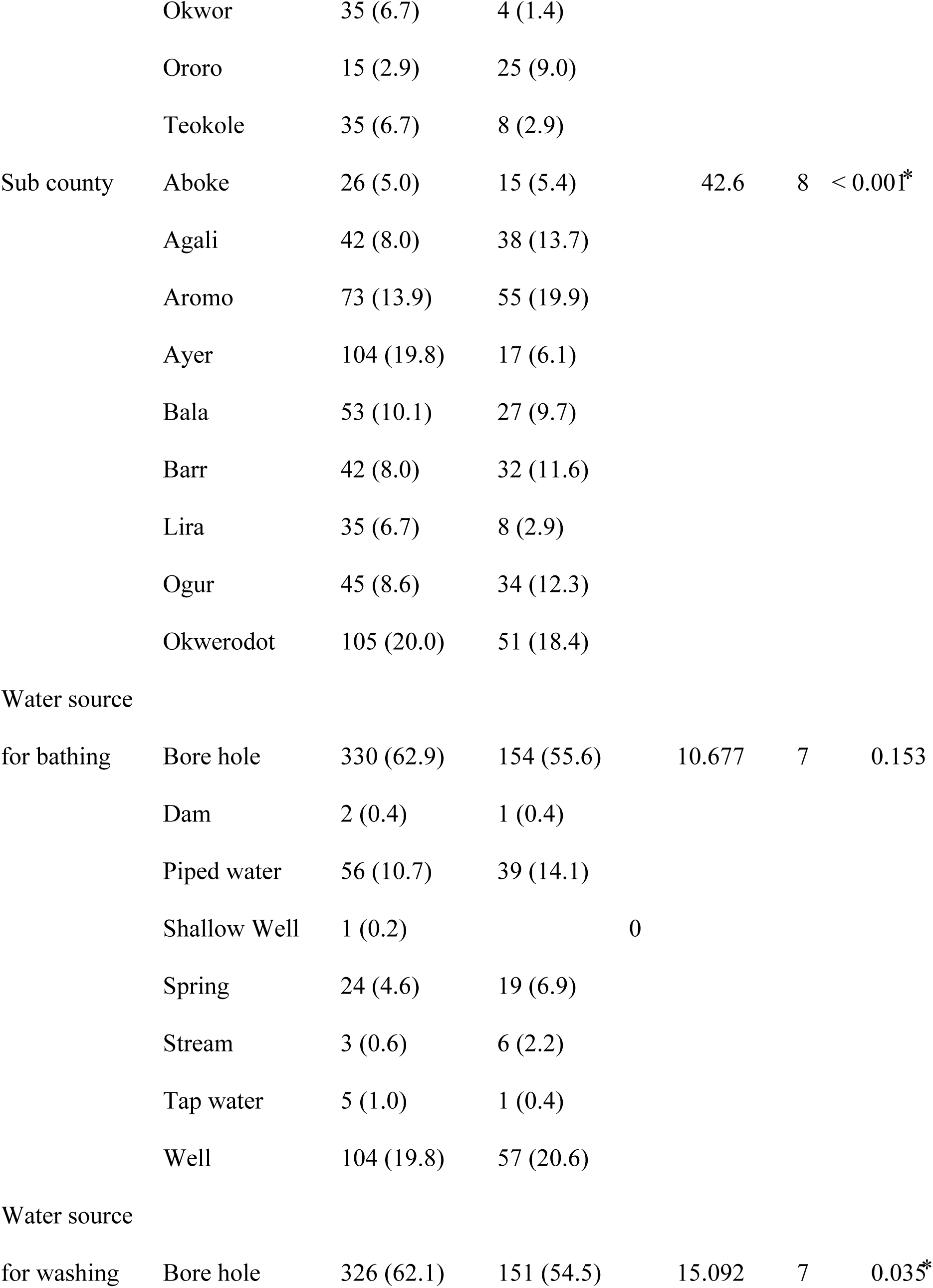
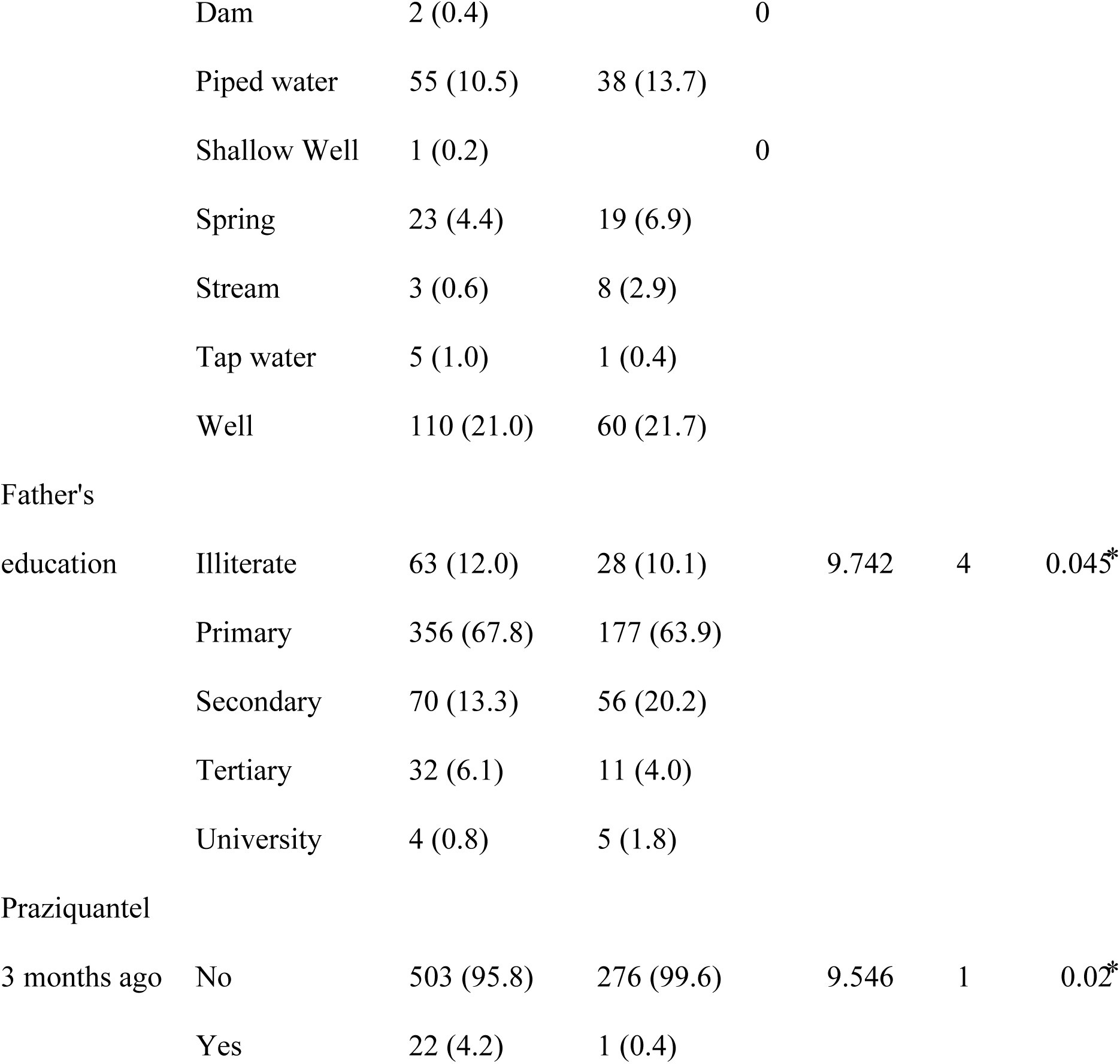
Prevalence of Schistosoma mansoni among school children.

The highest prevalence was in the 11-13 age group 154 (55.6%), but no significant association was found (p = 0.259). Participants in the P3 and P4 classes had the highest prevalence 79 (28.5%) each, while P2 and P1 had the lowest (p < 0.001). The sub-counties of Aromo 55 (19.9%) and Okwerodot 51 (18.4%) recorded the highest prevalence (p < 0.001), (Table 3).

There was no significant association between water source for bathing and infection status (p = 0.153). Significant differences were observed based on water sources for washing (p = 0.035), with streams 08 (2.9%) and wells 60 (21.7%) linked to higher prevalence. Lower prevalence was observed among participants with fathers having tertiary or university education (p = 0.045), while participants who received praziquantel within the last three months had significantly lower prevalence (p = 0.02), (Table 3).

### Intensity of Infection with *Schistosoma mansoni* among the Study Participants

The study assessed the infection intensity of *Schistosoma mansoni* infection among participants using eggs per gram (epg) of stool, categorized as light (1-99 epg), moderate (100-399 epg), and heavy (>400 epg). Overall infection intensity was light 93(11.6%), moderate 43(5.4%), and heavy 23(2.9%). In Kole District, 28 (7.04%) had light infections, 23 (5.78%) moderate, and 9 (2.26%) heavy infections. Lira City recorded only light infections 6 (13.95%) with no moderate or heavy cases. In Lira District, 59 (16.34%) had light infections, 20 (5.54%) moderate, and 14 (3.88%) heavy, (Table 4). Among females, 39 (10.80%) had light infections, 16 (4.43%) moderate, and 7 (1.94%) heavy.

**Table 4:**
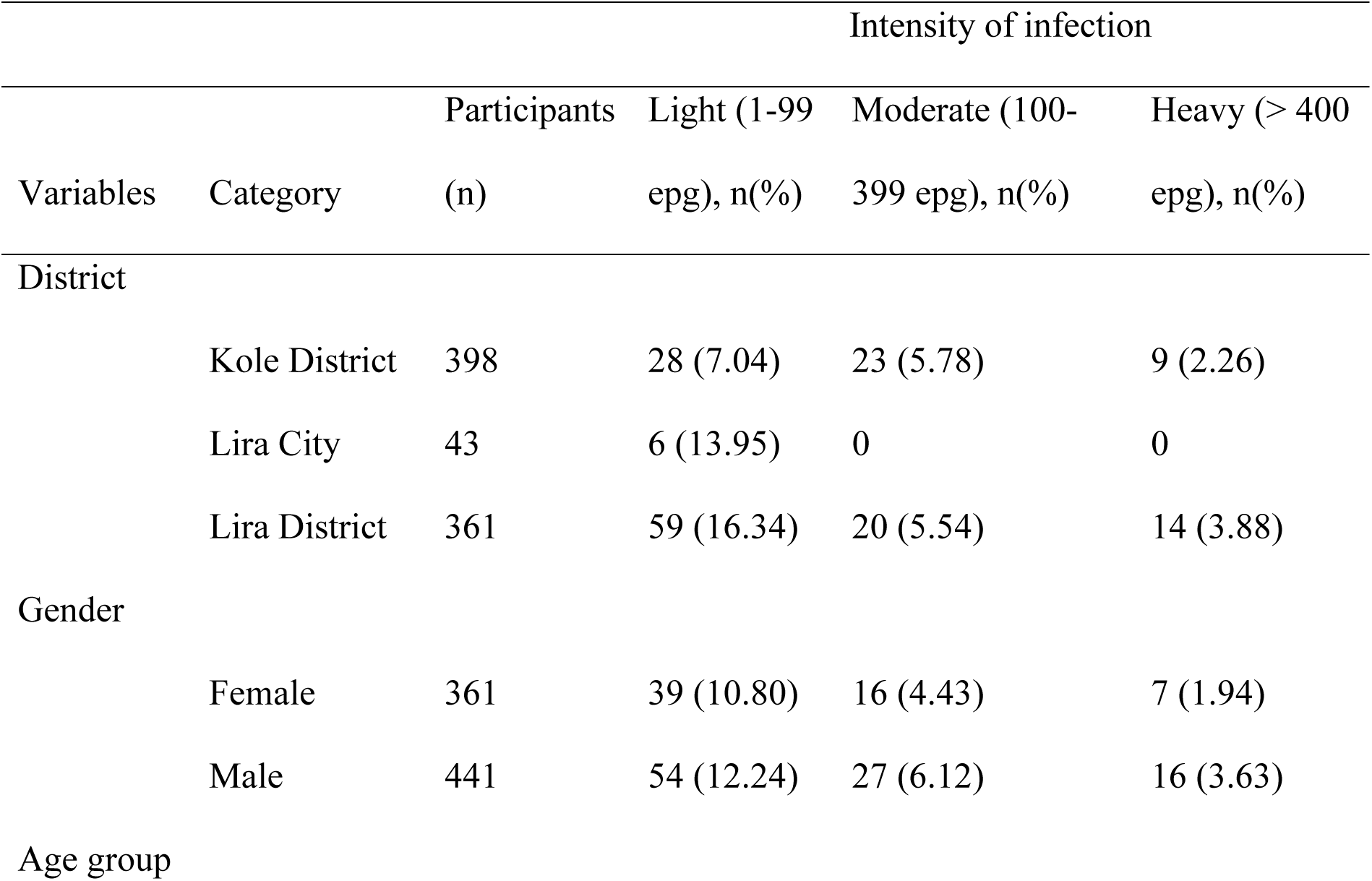
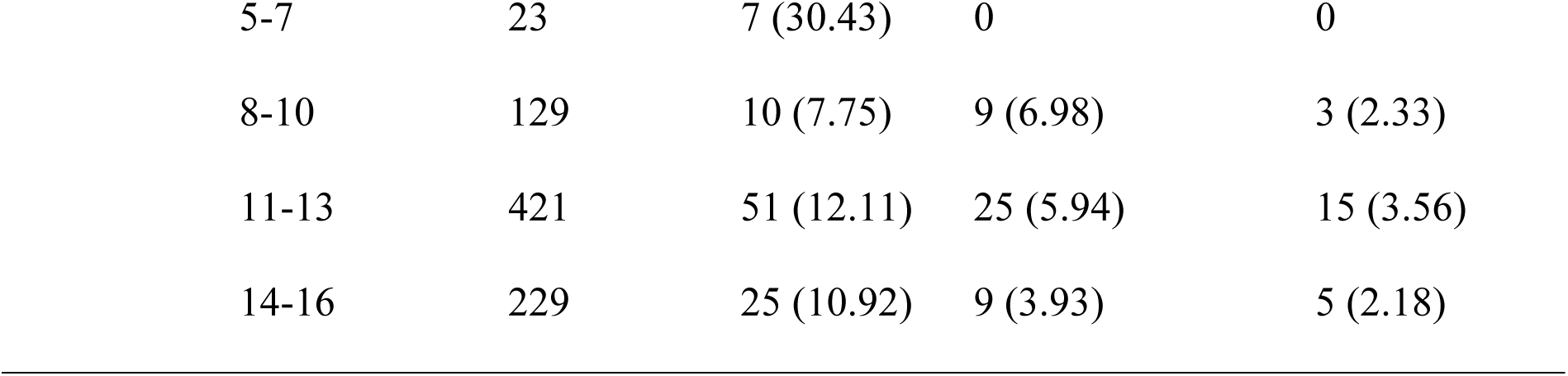
intensity of infection with *Schistosoma mansoni* among study participants.

|  |  | Intensity of infection |  |  |  |
| --- | --- | --- | --- | --- | --- |
| Variables | Category | Participants (n) | Light (1-99 epg), n(%) | Moderate (100-399 epg), n(%) | Heavy (> 400 epg), n(%) |
| District |  |  |  |  |  |
|  | Kole District | 398 | 28 (7.04) | 23 (5.78) | 9 (2.26) |
|  | Lira City | 43 | 6 (13.95) | 0 | 0 |
|  | Lira District | 361 | 59 (16.34) | 20 (5.54) | 14 (3.88) |
| Gender |  |  |  |  |  |
|  | Female | 361 | 39 (10.80) | 16 (4.43) | 7 (1.94) |
|  | Male | 441 | 54 (12.24) | 27 (6.12) | 16 (3.63) |
| Age group |  |  |  |  |  |

|  |  |  |  |  |
| --- | --- | --- | --- | --- |
| 5-7 | 23 | 7 (30.43) | 0 | 0 |
| 8-10 | 129 | 10 (7.75) | 9 (6.98) | 3 (2.33) |
| 11-13 | 421 | 51 (12.11) | 25 (5.94) | 15 (3.56) |
| 14-16 | 229 | 25 (10.92) | 9 (3.93) | 5 (2.18) |

Among males, 54 (12.24%) had light infections, 27 (6.12%) moderate, and 16 (3.63%) heavy. The 5-7 age group had the highest proportion of light infections 7 (30.43%) and no moderate or heavy infections. The 8-10 age group had 10 (7.75%) light, 9 (6.98%) moderate, and 3 (2.33%) heavy infections. The 11-13 age group had the highest proportions of moderate 25 (5.94%) and heavy infections 15 (3.56%), with 51 (12.11%) light infections. The 14-16 age group had 25 (10.92%) light, 9 (3.93%) moderate, and 5 (2.18%) heavy infections, (Table 4).

### Logistic regression analysis of risk factors for *Schistosoma mansoni* Infection

The logistic regression analysis identified several variables associated with *Schistosoma mansoni* infection. Participants from Kole District (OR = 0.038, 95% CI:0.009 - 0.162, p < 0.001) and Lira City (OR = 0.131, 95% CI:0.041-0.424, p = 0.001) had significantly lower odds of infection compared to those in Lira District, (Table 5).

**Table 5:** Logistic regression to determine risk factors associated with *Schistosoma mansoni* infection.

| Variables | Odd ratio | 95% C.I. |  | p-value |
| --- | --- | --- | --- | --- |
|  | (OR) | Lower | Upper |  |
| **District** |  |  |  |  |
| Kole District | 0.038 | 0.009 | 0.162 | < 0.001* |
| Lira City | 0.131 | 0.041 | 0.424 | 0.001* |
| Lira District | 1 | 1 | 1 | 1.000 |
| **School** |  |  |  |  |
| Aberdyangotoo | 6.219 | 1.48 | 26.138 | 0.013* |
| Abilonino | 0.624 | 0.106 | 3.675 | 0.602 |
| Abolet | 0.973 | 0.337 | 2.808 | 0.96 |
| Acutkumu | 0.122 | 0.042 | 0.351 | < 0.001* |
| Akano | 0.646 | 0.221 | 1.888 | 0.425 |
| Akore | 0.591 | 0.205 | 1.704 | 0.33 |
| Apedi | 7.558 | 1.886 | 30.291 | 0.004* |
| Ayara | 55.677 | 13.505 | 229.534 | < 0.001* |
| Ayer | 2.384 | 0.579 | 9.818 | 0.229 |
| Bala | 4.267 | 1.158 | 15.718 | 0.029* |
| Lwala | 2.247 | 0.533 | 9.462 | 0.27 |
| Obot | 0.147 | 0.044 | 0.487 | 0.002* |
| Obuto | 0.356 | 0.045 | 2.784 | 0.325 |
| Ocamoyang | 0.263 | 0.092 | 0.75 | 0.013* |
| Ogur | 0.307 | 0.096 | 0.982 | 0.046* |
| Oketkwer | 0.194 | 0.059 | 0.634 | 0.007* |
| Okwerodot | 3.911 | 0.948 | 16.13 | 0.059 |
| **Water source for bathing** |  |  |  |  |
| Borehole | 6.017 | 1.341 | 27.007 | 0.019* |
| Dam | 1.64409E<br>+11 | 0 | 0 | 0.999 |
| Piped water | 15.44 | 0.482 | 494.962 | 0.122 |
| Shallow well | 0 | 0 | 0 | 1.000 |
| Spring | 0 | 0 | 0 | 1.000 |
| Stream | 0 | 0 | 0 | 0.999 |
| **Water source for washing** |  |  |  |  |
| Borehole | 0.122 | 0.025 | 0.589 | 0.009* |
| Dam | 0 | 0 | 0 | 0.999 |
| Piped water | 0.077 | 0.002 | 2.591 | 0.153 |
| Spring | 25769101<br>40 | 0 | 0 | 1.000 |
| Stream | 26359246<br>8.9 | 0 | 0 | 0.999 |
| **Father's education** |  |  |  |  |
| Illiterate | 0.416 | 0.068 | 2.555 | 0.343 |
| Primary | 0.257 | 0.046 | 1.451 | 0.124 |
| Secondary | 0.4 | 0.074 | 2.167 | 0.288 |
| Tertiary | 0.179 | 0.032 | 1.003 | 0.05* |

Certain schools had significantly higher odds of infection, including Aberdyangotoo (OR = 6.219, p = 0.013), Apedi (OR = 7.558, p = 0.004), Ayara (OR = 55.677, p < 0.001), and Bala (OR = 4.267, p = 0.029), (Table 5). Conversely, schools such as Acutkumu (OR = 0.122, p < 0.001), Obot (OR = 0.147, p = 0.002), Ocamoyang (OR = 0.263, p = 0.013), Ogur (OR = 0.307, p = 0.046), and Oketkwer (OR = 0.194, p = 0.007) showed significantly reduced odds of infection, (Table 5).

Bathing with borehole water was significantly associated with higher odds of infection (OR = 6.017, p = 0.019). Washing with borehole water was protective (OR = 0.122, p = 0.009). Participants with fathers having tertiary education showed a borderline protective effect against infection (OR = 0.179, p = 0.05), (Table 5).

## Discussion

The findings of this study highlight the significant burden of *Schistosoma mansoni* and other intestinal parasitic infections among primary school children in the Lango sub-region of Northern Uganda. The observed prevalence of 277 (34.5%) for *S. mansoni* aligns with other studies conducted in endemic areas of Uganda [8] and sub-Saharan Africa [32], which report prevalence rates ranging from 20% to 60%, depending on geographical, environmental, and socio-economic factors [8, 12, 24, 32–34].

### Prevalence and Intensity of *Schistosoma mansoni* Infection

The high prevalence and intensity of *S. mansoni* infection in Lira District 159 (57.4%), compared to Kole District 110 (39.7%) and Lira City 8 (2.9%), suggest localized transmission hotspots that require targeted interventions [36]. The distribution of infections may be attributed to environmental factors, such as the presence of freshwater bodies, where children frequently engage in activities such as bathing, swimming, and fetching water. Prior studies indicate that open-water contact is a major risk factor for schistosomiasis transmission, particularly in regions where hygienic water sources are scarce [35–38]. The lower prevalence in Lira City may reflect better access to piped water, improved sanitation, and urbanization-related factors that reduce exposure risks [40].

The observed infection intensities, light 93 (11.6%), moderate 43 (5.4%), and heavy 23 (2.9%) underscore the ongoing transmission of schistosomiasis in this population. Similar studies have reported variations in infection intensities based on factors such as age, gender, and previous treatment history [12, 24, 33, 39]. The presence of moderate and heavy infections indicates a high parasite burden, necessitating urgent deworming interventions and reinforcement of preventive strategies [42].

### Risk factors associated with Schistosomiasis

The study identified several significant risk factors for *S. mansoni* infection. Notably, bathing with borehole water was significantly associated with higher odds of infection (OR = 6.017, p = 0.019). This suggests that borehole water may not be entirely safe, either due to contamination with cercariae or poor water storage practices [43]. In contrast, washing with borehole water was found to be protective (OR = 0.122, p = 0.009), which aligns with findings that suggest that schistosome transmission is more strongly associated with prolonged skin contact in contaminated water bodies rather than routine household washing [24, 40–42].

Parental education level also influenced infection rates. Children whose fathers had a tertiary education had a lower risk of infection (OR = 0.179, p = 0.05), indicating that higher education levels are associated with better knowledge of hygiene and preventive health behaviors [47].

Similar findings have been reported in other schistosomiasis-endemic regions, emphasizing the role of socioeconomic factors in disease prevention [39, 43–45]. Households with higher education levels may also have better access to clean water, sanitation, and preventive healthcare services, further reducing infection risks [51].

The study also found that recent praziquantel treatment significantly reduced schistosomiasis prevalence (OR = 0.122, p = 0.009), reinforcing the importance of Mass Drug Administration (MDA) programs in endemic settings. However, despite MDA campaigns, the persistence of moderate and heavy infections suggests potential gaps in treatment coverage, compliance, or reinfection rates [46–50]. Reinfection often occurs due to continued exposure to contaminated water sources, underscoring the need for sustained health education, environmental sanitation, and behavioral change interventions to complement chemotherapy programs [51, 52].

### Co-infection with other intestinal parasites

The study also identified the presence of intestinal parasites, with *Entamoeba coli* 94 (11.7%) and *Ascaris lumbricoides* 93 (11.6%) being the most prevalent. The occurrence of soil-transmitted helminths (STHs), including *Hookworms* 51 (6.4%) and *Trichuris trichiura* 12 (1.5%), further highlights the burden of neglected tropical diseases (NTDs) in the study area [53–55]. The co-existence of schistosomiasis and STHs has been reported in several regions, particularly where poor sanitation and hygiene (WASH) conditions prevail [56–58]. Such co-infections contribute to malnutrition, anemia, and cognitive impairments, necessitating integrated control strategies that address both waterborne and soil-transmitted parasitic infections [59–62].

The presence of rare infections, such as Fasciola species 11 (1.4%) and Taenia species 7 (0.9%), suggests possible zoonotic transmission, likely influenced by livestock interactions and consumption of contaminated food or water [39, 63]. These findings emphasize the need for a One Health approach to disease control, incorporating human, animal, and environmental health interventions [64, 65].

## Conclusion

The findings of this study underscore the significant burden of schistosomiasis and intestinal parasitic infections among primary school children in the Lango sub-region. The high prevalence and intensity of *S. mansoni* infections, particularly in the Lira District, highlight urgent public health concerns. Key risk factors, including bathing with borehole water, low parental education, and school-based exposure, indicate the need for targeted interventions beyond mass drug administration. Implementing integrated, community-driven strategies, including improved sanitation, health education, and environmental modifications, is essential to sustainable schistosomiasis control. Future research should focus on reinfection dynamics, treatment efficacy, and long-term control measures to guide policy formulation and intervention planning.

## Declarations

### Ethics approval and consent to participate

Ethical approval for this study was obtained from the Gulu University Research and Ethics Committee (GUREC-2022-323). Additional approval was obtained from the Uganda National Council for Science and Technology (UNCST-HS2571ES). Written informed consent was obtained from parents or legal guardians of participating children, and assent was obtained from children aged 8 years and above. All methods were performed in accordance with relevant guidelines and regulations.

### Consent for publication

Not applicable.

### Availability of data and materials

The datasets used and analyzed during the current study are available from the corresponding author upon reasonable request.

## Authors’ contributions

JPB conceptualized the study, collected and analyzed the data, and drafted the manuscript. RO, MN, GMM, RE, and EIOA contributed to the study design, data analysis, and manuscript review. All authors read and approved the final manuscript.

## Acknowledgments

The authors thank the participating schools, children, parents, and local authorities in Lira District, Lira City, and Kole District for their cooperation and support. Appreciation also goes to Gulu University for providing administrative and logistical support during the study.

